# Imaging synaptic microstructure and synaptic loss *in vivo* in early Alzheimer’s Disease

**DOI:** 10.1101/2021.11.23.21266746

**Authors:** Ashwin V Venkataraman, Courtney Bishop, Ayla Mansur, Gaia Rizzo, Yvonne Lewis, Ece Kocagoncu, Anne Lingford-Hughes, Mickael Huiban, Jan Passchier, James B Rowe, Hideo Tsukada, David J. Brooks, Laurent Martarello, Robert A. Comley, Laigao Chen, Richard Hargreaves, Adam J. Schwarz, Roger N. Gunn, Eugenii A. Rabiner, Paul M Matthews

**Affiliations:** Department of Brain Sciences, Imperial College London; London, UK; UK Dementia Research Institute at Imperial College London; London, UK; Invicro LLC.; London, UK; King’s College London; London, UK; University of Cambridge; Cambridge, UK; Hamamatsu Photonics; Hamamatsu,Japan; University of Newcastle upon Tyne, UK and Aarhus University; DK; Biogen, Cambridge; USA; AbbVie, Chicago; USA; Pfizer Inc., Cambridge; USA; Bristol Myers Squibb; New York, USA; Takeda Pharmaceuticals Ltd; Cambridge, USA; MINDMAPS Consortium

**Keywords:** Synaptic microstructure, synaptic loss, imaging, Alzheimer’s, NODDI, free water, SV2A

## Abstract

1.

**Background:** Synaptic loss and neurite dystrophy are early events in Alzheimer’s Disease (AD). We aimed to characterise early synaptic microstructural changes *in vivo*.

**Methods:** MRI neurite orientation dispersion and density imaging (NODDI) and diffusion tensor imaging (DTI) were used to image cortical microstructure in both sporadic, late onset, amyloid PET positive AD patients and healthy controls (total n = 28). We derived NODDI measures of grey matter extracellular free water (FISO), neurite density (NDI) and orientation dispersion (ODI), which provides an index of neurite branching and orientation, as well as more conventional DTI measures of fractional anisotropy (FA), mean/axial/radial diffusivity (MD, AD, RD, respectively). We also performed [^11^C]UCB-J PET, which provides a specific measure of the density of pre-synaptic vesicular protein SV2A. Both sets of measures were compared to regional brain volumes.

**Results:** The AD patients showed expected relative decreases in regional brain volumes (range, -6 to - 23%) and regional [^11^C]UCB-J densities (range, -2 to -25%). Differences between AD and controls were greatest in the hippocampus. NODDI microstructural measures showed greater FISO (range, +26 to +44%) in AD, with little difference in NDI (range, -1 to +7%) and mild focal changes in ODI (range, -4 to +3%). Regionally greater FISO and lower [^11^C]UCB-J binding were correlated across grey matter in patients (most strongly in the caudate, r^2^ = 0.37, p = 0.001). FISO and DTI RD were strongly positively associated, particularly in the hippocampus (r^2^ = 0.98, p < 7.4 × 10^−9^). After 12-18 months we found a 5% increase in FISO in the temporal lobe, but little change across all ROIs in NDI and ODI. An exploratory analysis showed higher parietal lobe FISO was associated with lower language scores in people with AD.

**Conclusions:** We interpreted the increased extracellular free water as a possible consequence of glial activation. The dynamic range of disease-associated differences and the feasibility of measuring FISO on commercially available imaging systems makes it a potential surrogate for pathology related to synapse loss that could be used to support early-stage evaluations of novel therapeutics for AD.

## BACKGROUND

Alzheimer’s Disease (AD) is an ultimately fatal brain disease associated with progressive cognitive, social and functional decline. AD affects 50 million people worldwide. [1] The number of people affected will triple by 2050 without effective prevention strategies or treatments.[2]

AD begins with biochemical pathology, neuroinflammatory responses involving both microglial and astrocyte activation, and the deposition of amyloid-β (Aβ) [3–5] and tau aggregates.[6,7] Whilst substantial progress has been made in defining the protein pathology and the clinical phenotype, the pathogenesis, timing and distribution of early synaptic microstructural changes in humans are still poorly understood.

Most knowledge of synaptic pathology in AD has emerged from preclinical models or studies of *post mortem* human brain tissue. Clinical-pathological correlations suggest a strong relationship between loss of synaptic integrity and cognitive dysfunction [8,9]. One model of AD pathogenesis proposes that amyloid oligomers induce maladaptive microglial synaptic pruning that leads to dendritic and axonal degeneration. [10–12] This loss of neuropil is believed to be a major contributor to the loss of brain volume as the disease progresses[13,14].

*In vivo* characterisation of early microstructural changes in AD complements neuropathological studies of *post mortem* human tissues and AD-related preclinical models. Informative and clinically practical measures of microstructural changes and synaptic loss promises earlier diagnosis and improved monitoring of disease progression and responses to new disease modifying therapies. [15]

Neurite Orientation Dispersion and Density Imaging (NODDI) is an advanced approach to *in vivo* diffusion-weighted MRI based on a multi-compartment diffusion model which allows a more precise characterisation of water diffusion than diffusion tensor imaging (DTI). [16] When applied to brain grey matter, NODDI provides measures related to the *extra*cellular free water volume fraction (FISO), neurite density (NDI) (the *intra*cellular volume fraction), and the local diffusion tensor orientation dispersion (ODI) (interpretated in this context as a measure of neurite branching). [^11^C]UCB-J PET is a novel PET radioligand targeting the pre-synaptic vesicle glycoprotein 2A (SV2A) that allows the assessment of synaptic density *in vivo* in humans [17] [18] [19]. Here we used NODDI MRI and [^11^C]UCB-J PET methods together to assess the microstructural pathology in AD and to explore its relationship to measures of clinical disease expression. We studied patients with sporadic, late onset AD and a comparable healthy control population.

## METHODS

### Study Participants

12 patients with typical amnestic early AD were recruited for this study. Previously acquired data from 16 healthy controls in the MINDMAPS healthy volunteers cohort was used as a control group. [20] All participants underwent screening, [^11^C]UCB-J dynamic PET imaging, and volumetric and NODDI MRI scanning. AD patients underwent repeated assessments and scanning 12-18 months after their initial (baseline) scans.

AD patients met National Institute on Aging-Alzheimer’s Association (NIA-AA) core clinical criteria for probable AD dementia or amnestic mild cognitive impairment. All were Aβ positive based on [^18^F]Florbetaben PET imaging (see below), aged >50 with a Mini Mental State Examination (MMSE) score ≥18 and able to give informed consent to participate in the study. Patients taking symptomatic therapy for AD had been on a stable dose for at least 6 weeks prior to their baseline evaluation.

Exclusion criteria for both groups included histories of major psychiatric, neurological or medical illnesses or current infections, use of a medication known to either bind directly to SV2A (e.g., levetiracetam) or impair cognition (e.g., opiates, anticholinergics, benzodiazepines) or exposure to >10mSv of ionising radiation in the past year. Routine clinical blood haematological and biochemical tests and *APOE* genotyping were performed. Neuropsychological and cognitive assessments included the Mini Mental State Examination (MMSE), Addenbrooke’s Cognitive Exam (ACE-III), Repeatable Battery for Assessment of Neuropsychological Status (RBANS) and the National Adult Reading Test (NART). Specific RBANS tests included assessing immediate memory (list learning and story memory), visuospatial perception (figure copy and line orientation), language (picture naming and semantic fluency), attention (digit span and coding) and delayed memory (list recall, list recognition, story memory and figure recall). Specific ACE-III tests included rating attention (orientation), verbal memory (word recall, address recall, semantic recall), fluency (word generation), language (sequenced commands, writing, repetition, pronunciation) and visuospatial memory (figure copy, clock drawing, dot counting, letter recognition).

All participants provided written informed consent. Ethical approval for this study was provided by the NHS London - Brighton & Sussex Research Ethics Committee (REC 18/LO/0179) for AD patients and East of England Cambridge Central & South Research Ethics Committee for controls. Radiation safety was approved by the Administration of Radioactive Substances Advisory Committee (ARSAC R92), London. Local site approval was provided by Imperial College London Joint Research Office. Participants were recruited from Imperial College Healthcare NHS Trust, West London Mental Health Trust, and Cambridge University Hospitals NHS Trust. All screening and scanning were conducted at the Invicro imaging centre, Hammersmith Hospital Site, London, UK.

### Image Acquisition

[^11^C]UCB-J was synthesised and PET/CT images and arterial blood samples were acquired as previously described[18,21]. [^18^F]Florbetaben was purchased from Alliance Medical. Imaging was performed using either a Siemens Biograph 6 True Point or HiRez PET/CT scanner (Siemens Healthcare, Erlangen, Germany). A low-dose CT scan was acquired to enable attenuation correction (30 mAs, 130 KeV, 0.55 pitch) prior to each imaging session. AD patients received a [^18^F]Florbetaben scan at screening which was acquired over 20 min following a 90 min distribution phase. [^11^C]UCB-J was administered as an intravenous bolus (20 ml over 20 seconds), and serial blood samples were collected from the radial artery to assess changes in the concentration of radioactivity in whole blood and plasma, as well as the percent of unmetabolized tracer over the course of the scan. Following tracer administration, dynamic emission data were acquired in list mode over 90 minutes and reconstructed into 26 frames (frame durations: 8×15 s, 3×60 s, 5×120 s, 5×300 s, 5×600 s) using discrete inverse Fourier transform reconstruction with corrections for attenuation, random coincidences and scatter.

All MRIs were acquired using a Siemens 3T Trio clinical scanner (Siemens Healthineers, Erlangen, Germany) with a 32-channel phased-array head coil. T1-weighted structural data was acquired using a 3D MPRAGE sequence (TE = 2.98 ms, TR = 2300 ms, flip angle = 9°, voxel size = 1.0 mm x 1.0 mm x 1.0 mm). Diffusion Tensor Imaging (DTI) was acquired with 64 diffusion directions at b = 1000 plus six b = 0 scans interspersed, using 3 x acceleration (MB = 3), two runs to provide acquisition in both the positive and negative phase-encoding direction and TE = 88.0 ms, TR = 3010 ms and a nominal voxel size = 1.9×1.9×2.0 mm. Neurite Orientation Dispersion and Density Imaging (NODDI) was performed using 2-shells with 32 directions at b=700 (plus 4 b=0’s) and 64 directions at b=2000 (plus 8 b=0’s) and TE = 101.8 ms, TR = 3340 ms, MB = 3 and a nominal voxel size 1.9×1.9×2.0 mm.

### Image and statistical analyses

[^18^F]Florbetaben PET data were quantified by Amyloid IQ[22], using 33% as the cut-off for amyloid positivity and inclusion in the study. The [^11^C]UCB-J PET image analysis pipeline, which included generation of an arterial input function, tracer kinetic modelling, model comparison and selection and time stability assessment, has been described previously[20]. PET data were processed using MIAKAT™ (version 4.3.24, miakat.org). MIAKAT™ is implemented using MATLAB (version R2016a; Mathworks) and makes use of SPM12 (Wellcome Trust Centre for Neuroimaging, fil.ion.ucl.ac.uk/spm) functions for image segmentation and registration. Individual participant structural MRIs underwent grey matter segmentation and rigid body co-registration to a standard reference space.[23] The template brain image and associated CIC neuroanatomical atlas then was nonlinearly warped to the individual participant’s MRI, for which the regions of interest (ROI) were defined and volumes of regions in mm^3^ were measured. A centrum semiovale (CS) ROI also was generated from the automated anatomic labelling template as defined previously for use as a non-specific reference region for [^11^C]UCB-J.[18] PET images were registered to each participant’s MRI and corrected for motion using frame-to-frame rigid-body registration. Regional time activity curves (TAC) were generated for each ROI.

Regional volumes of distribution (V_T_) were calculated using the one-tissue compartmental model (1TC) for [^11^C]UCB-J as described previously[20]. Partial volume-corrected data were generated using the Muller-Gartner algorithm.[24] The outcome parameter chosen for [^11^C]UCB-J was the DVR_CS-1_, that uses the CS as a pseudo-reference region in which

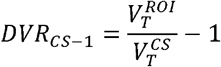

with 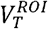 describes the partial volume corrected V_T_ in the region of interest and 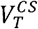 describes the V_T_ in the centrum semiovale.

Comparison of regional target binding between AD patients and control groups was performed using a two-tailed, unpaired Student’s t-test for cross-sectional data and a two-tailed, paired Student’s t-test for longitudinal data across *a priori* selected regions (frontal lobe, parietal lobe, temporal lobe, cerebellum, hippocampus, thalamus, caudate, anterior cingulate and posterior cingulate).

For DTI and NODDI processing, T1w data was initially processed using FSL (*fsl_anat*) to provide tissue segmentations for the individual participant’s T1w image (fsl.fmrib.ox.ac.uk)[25]. GM and WM PVE maps were thresholded at 0.5 and binarized to provide GM and WM definitions, respectively. The CIC Atlas [23] was nonlinearly aligned to the subject’s native T1w image to provide native-space ROIs.

For DTI processing, we estimated the susceptibility-induced off-resonance field from B0s (with AP and PA phase encoding) using FSL’s *topup* tool, followed by correction of geometric distortion in the full DTI acquisition using FSL’s *<u>eddy</u>* tool. DTI parameters (FA, MD, L1, RD) were computed from eddy-corrected DTI data using FSL’s *dtifit* tool. Finally, FSL’s *epi_reg* tool provided the DTI-to-T1 alignment parameters, and the DTI parameter maps were transformed to the individual participant’s T1w image.

For NODDI processing, the DTI-to-NODDI (b0) alignment parameters were computed using FSL’s flirt tool, then geometric distortion correction of the NODDI data was performed using FSL’s eddy tool (providing DTI-to-NODDI transformation parameters for fieldmap estimated in NODDI space). Estimation of NODDI parameters (ficvf (referred to here as NDI), ODI, FISO) from the eddy-corrected NODDI data was achieved with the University College London Microstructure Imaging group NODDI MATLAB toolbox (http://mig.cs.ucl.ac.uk/index.php?n=Download.NODDI). Finally, computation of NODDI-to-T1 alignment parameters used FSL’s epi_reg tool and the NODDI parameter maps were aligned to the individual participant’s T1w image.

Pearson’s r correlations were performed across modalities and groups for NODDI measures (NDI, ODI, FISO), [^11^C]UCB-J and regional volumes. Group cross-sectional imaging data of AD patients compared to controls was computed as z-scores (AD patients mean – controls mean)/(controls standard deviation). Associations between imaging findings, neuropsychological and cognitive performance variables were explored. Correlations between these measures and neuropsychological and cognitive performance measures also were explored for *a priori* hypothesis generation. Corrections for multiple comparisons were calculated using the false discovery rate. All statistics were analysed and plotted using R software (R-project.org).

## RESULTS

### Subject characteristics

28 participants contributed data to this study. 12 participants had a diagnosis of sporadic onset amnestic, amyloid-positive early-stage AD and 16 were healthy controls (Table 1). One NODDI scan in each group could not be processed due to inability of the participant to complete the full scan, so 11 NODDI scans were available for assessment in the AD group and 15 for the controls. For DTI 11 scans were available in the AD group and 16 for controls. Both groups had similar premorbid intelligence as measured by the NART with equal distributions of males and females. The AD patients had a lower MMSE, immediate memory, language and delayed memory test scores than the controls (Table 1). 8 participants with early AD were able to complete both baseline and 12-18 month follow-up SV2A PET scans and 10 patients were able to complete follow-up structural and NODDI MRI scans.

**Table 1.**
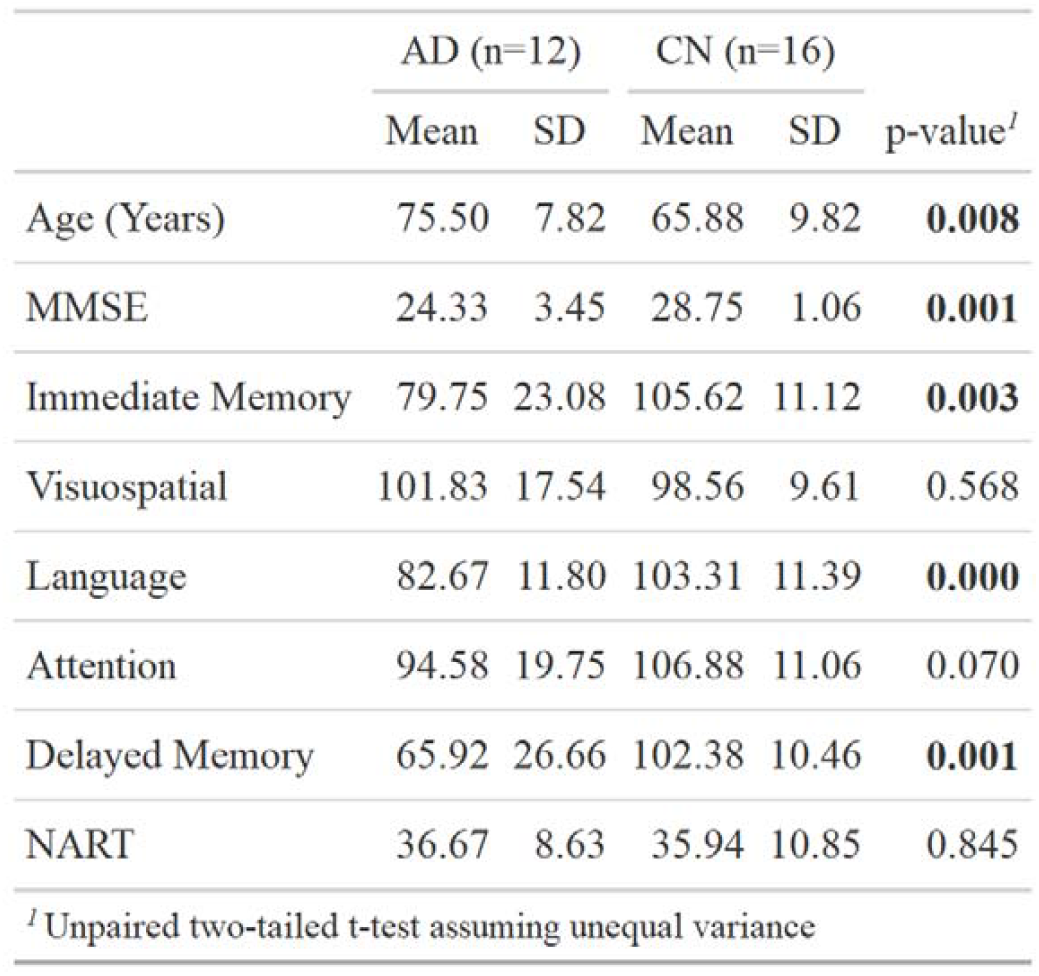
Cross-sectional demographic descriptive data and neuropsychological test results for AD patients and controls at baseline.

### Brain volume and synaptic density are both reduced in early AD

All AD patient brain ROIs had lower volumes than those of the healthy controls. The greatest relative differences were in the hippocampus (mean -23%, median -18%, range -48% to -2%, p < 0.001) and temporal lobe grey matter (mean -16%, median -18%, range -33 to +5%, p < 0.001) (Figure 1). [^11^C]UCB-J DVR_CS-1_ was lower in brains of the AD patients than in controls (Figure 2). The greatest relative differences in SV2A density were found in the caudate (mean -25%, median -28%, range -75 to +16%, p=0.007), hippocampus (mean -24%, median -23%, range -52 to +6%, p=0.001) and thalamus (mean -19%, median -20%, range -44 to +21%, p=0.012) (Figure 2).

**Figure 1.**
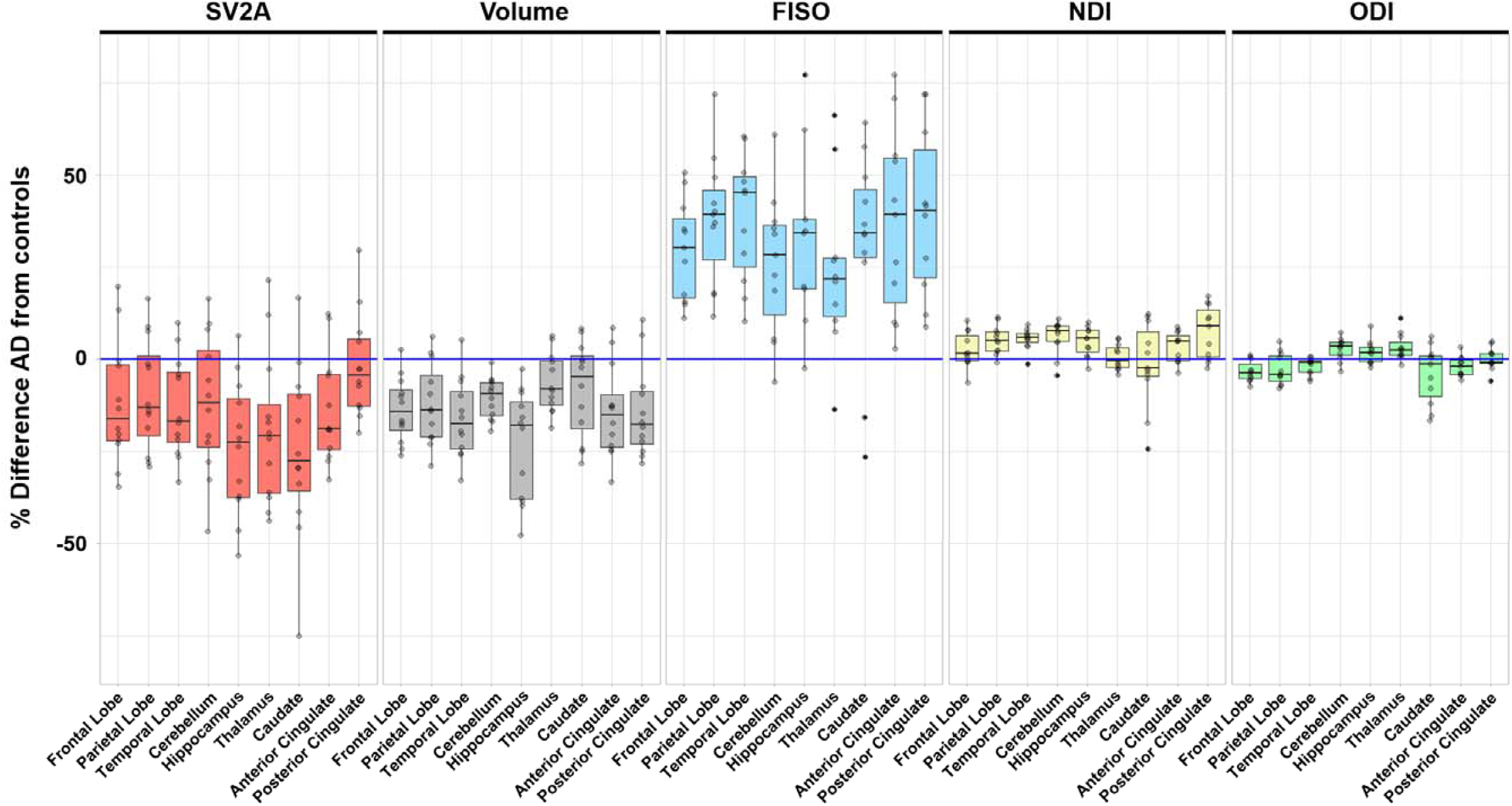
Cross-sectional percentage differences for individual AD patient (with group mean values and standard deviations) relative to the control mean for NDI, ODI, FISO, SV2A and normalised brain volume measures.

**Figure 2.**
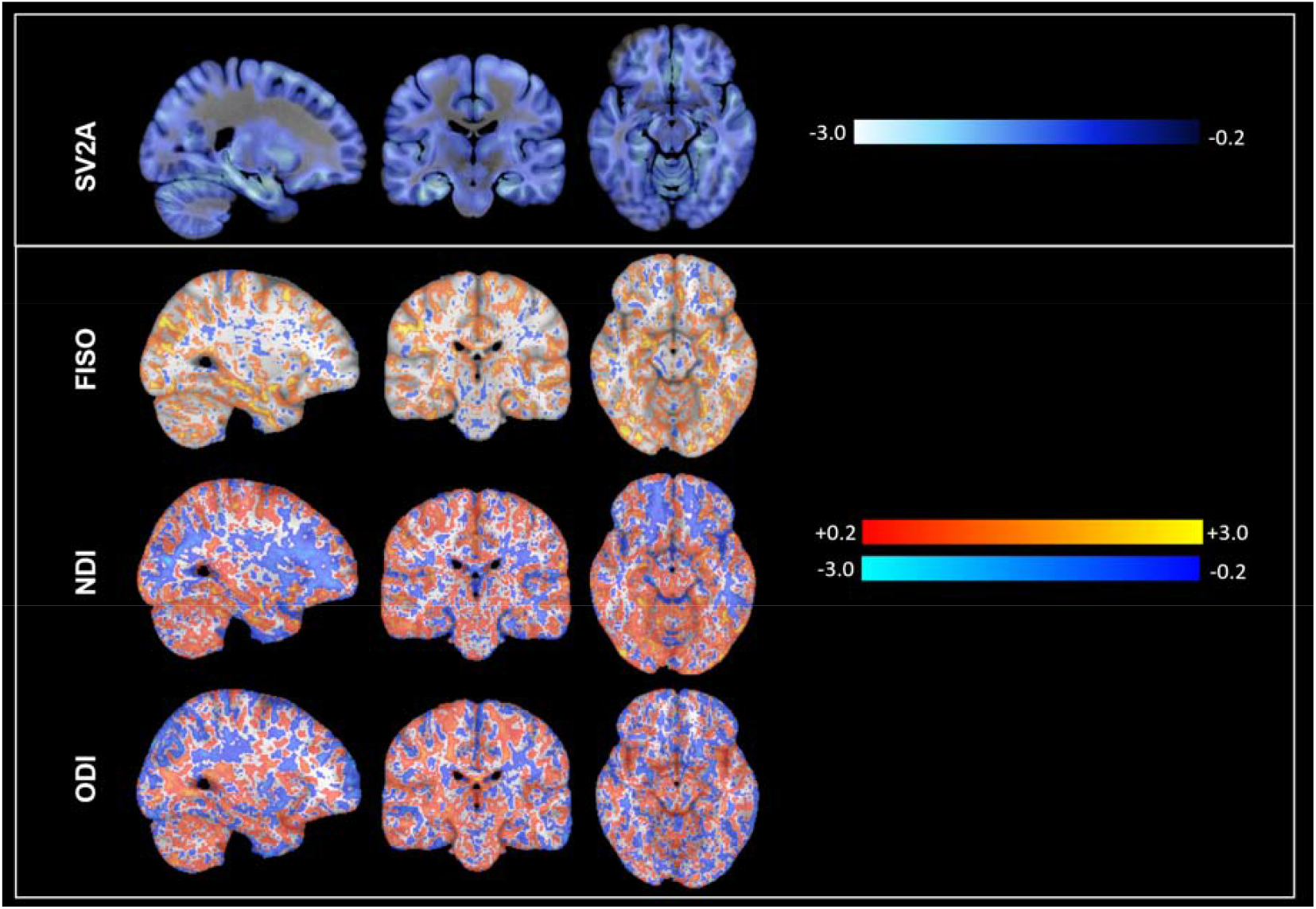
AD z score images on MNI152 for [^11^C]UCB-J DVR_CS-1_ PET (SV2A) and NODDI images of free water (FISO) neurite density (NDI) and orientation dispersion (ODI) (top of image to bottom). Spatially normalised AD patient images acquired at the study baseline were contrasted to those from the control group. Thresholded voxel-wise z-scores ((AD patients mean – controls mean)/(controls standard deviation)) are illustrated, using the group mean control image as a template. Z-score scales are shown to the right of the images.

### Cortical free water is increased in AD

NODDI FISO was higher in AD compared to the controls in all brain grey matter brain regions (range +26 to +44%) (Figure 1). The greatest relative differences in FISO were observed in the hippocampus (mean +44%, range -2 to 102%, p = 0.001) and posterior cingulate (mean +44%, range 9 – 91%, p < 0.001) (Figure 2). The strongest negative association between regional brain volume and FISO in the AD brains was found for the hippocampus (r^2^ = 0.88, p < 0.0001) (Figure 3).

**Figure 3.**
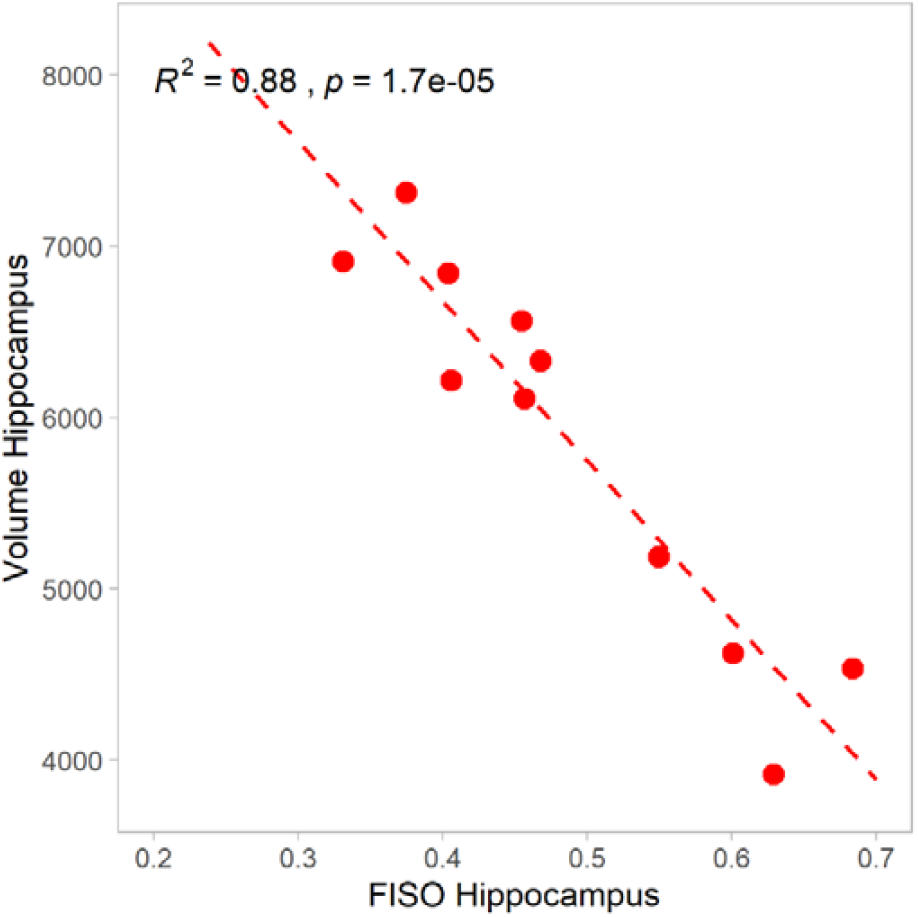
Highly significant negative associations between the normalised hippocampal volume and regional FISO in AD patients were found.

NDI values were also larger in the brains of the AD patients compared with the controls, but the relative differences were smaller than those observed with FISO (Figure 2). The greatest increases in NDI with AD were in the parietal (mean +5%, range -1 to +12%, p = 0.007) and temporal (mean +5%, range -1 to +9%, p < 0.001) lobes, cerebellum (mean +6%, range -4 to +10%, p = 0.002), hippocampus (mean +5%, range -3 to +10%, p = 0.045) and posterior cingulate (mean +7%, range -2 to +17%, p = 0.011).

The ODI values were nominally lower in the AD patient brains compared to controls for some ROIs, but the magnitudes of the differences were small (Figure 2). The largest differences were found in the frontal (mean -3%, range -8 to +1%, p = 0.007) and medial temporal lobes (mean -2%, range -6 to 0%, p = 0.038).

We compared cross-sectional effect sizes between AD patients and controls for FISO, SV2A PET, and regional volumes using Hedges g (Table 2). The effect sizes (Hedge’s g) were higher for FISO (range 0.82 - 2.30) than for the other NODDI and DTI measures in most ROIs and, notably, in the temporal lobe.

**Table 2.** Cross-sectional effect sizes using Hedges g for differences in SV2A PET, volume, NODDI FISO, NDI, ODI, and DTI FA, MD, L1 and RD measures for the AD patients compared to controls.

| ROI | Effect sizes (Hedges g) |  |  |  |  |  |  |  |  |
| --- | --- | --- | --- | --- | --- | --- | --- | --- | --- |
|  | SV2A | Volume | FISO | NDI | ODI | FA | MD | L1 | RD |
| Frontal Lobe | -0.80 | -1.33 | 1.67 | 0.54 | -1.19 | 0.43 | 1.50 | 1.58 | 1.43 |
| Parietal Lobe | -0.69 | -1.19 | 1.82 | 1.09 | -0.65 | 0.20 | 1.69 | 1.69 | 1.69 |
| Temporal Lobe | -1.12 | -1.54 | 2.30 | 1.70 | -0.89 | -0.54 | 1.89 | 1.95 | 1.86 |
| Cerebellum | -0.82 | -1.60 | 1.35 | 1.44 | 0.74 | -0.69 | 1.43 | 1.48 | 1.39 |
| Hippocampus | -1.52 | -1.97 | 1.66 | 0.75 | 0.55 | -1.18 | 1.37 | 1.38 | 1.37 |
| Thalamus | -1.07 | -0.80 | 1.05 | 0.09 | 0.64 | -1.00 | 0.77 | 0.74 | 0.78 |
| Caudate | -1.17 | -0.60 | 0.82 | -0.13 | -0.49 | -1.12 | 0.69 | 0.61 | 0.73 |
| Anterior Cingulate | -1.06 | -1.16 | 1.54 | 0.66 | -0.76 | -0.22 | 1.32 | 1.36 | 1.29 |
| Posterior Cingulate | -0.16 | -1.16 | 1.66 | 1.12 | 0.02 | -0.15 | 1.35 | 1.42 | 1.30 |

### Lower SV2A synaptic density correlates strongly with increased grey matter extracellular free water

We tested for relationships between SV2A and NODDI measures (Figure 4). We found negative correlations between SV2A and FISO in AD across multiple ROIs, particularly in the parietal lobe (r^2^ = 0.47, p < 0.02), and for both AD patients and controls in the caudate (r^2^ = 0.37, p < 0.001) and parietal lobe (r^2^ = 0.21, p = 0.018).

**Figure 4.**
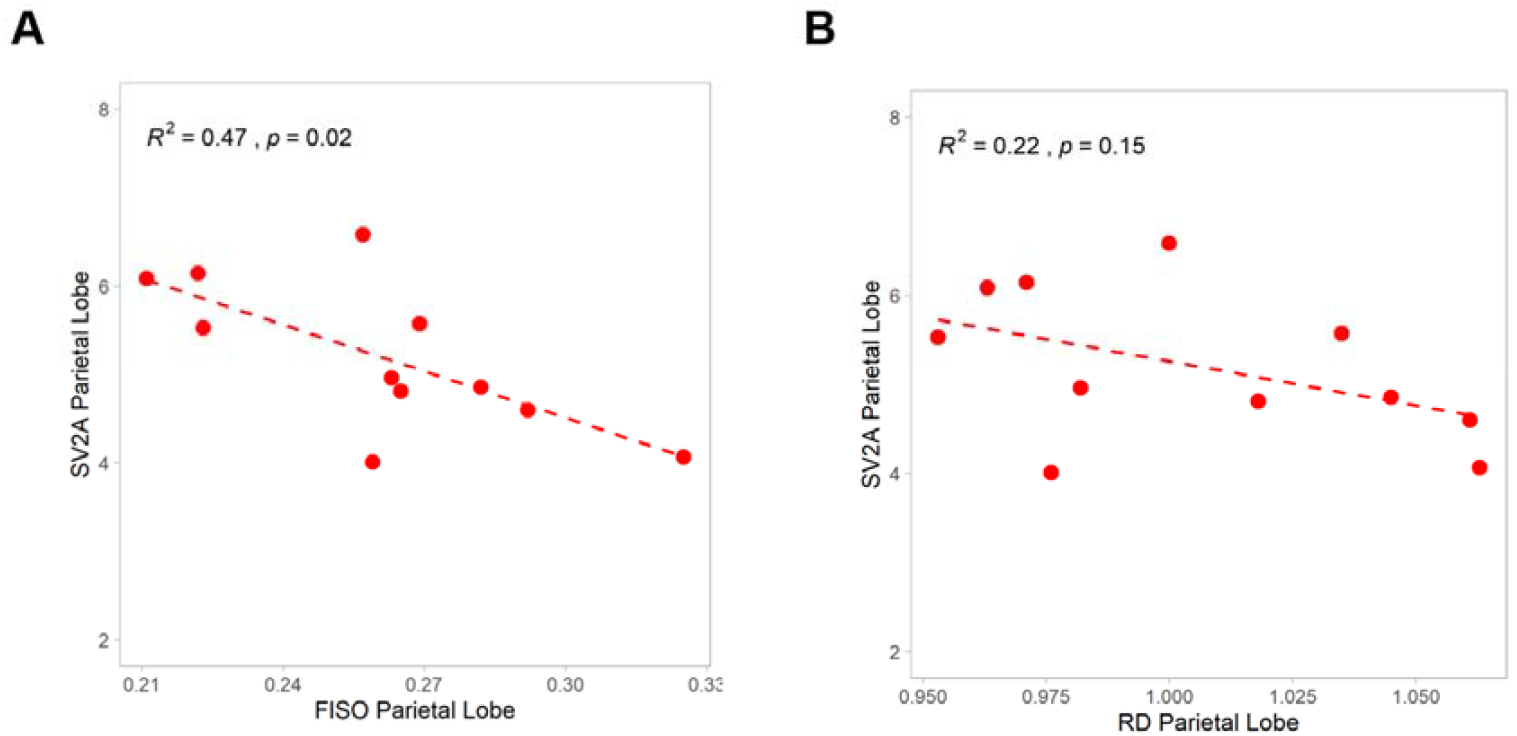
A negative association between SV2A and FISO (A) and a trend to a negative association for SV2A and RD (B) in the parietal lobe in AD were found.

Weaker correlations for AD and controls were found between SV2A and NDI, but these were present only for a subset of the grey matter ROIs, such as the frontal (r^2^ = 0.17, p < 0.041) and parietal (r^2^ = 0.33, p < 0.0028) lobes (but not, e.g., in the hippocampus, r^2^ = 0.022, p = 0.47). Similarly, SV2A and ODI were weakly correlated in only some ROIs (e.g., the parietal lobe, r^2^ = 0.15, p < 0.049 and caudate, r^2^ = 0.2, p < 0.025).

### Higher DTI RD correlates strongly with NODDI FISO in brain grey matter

DTI RD was globally significantly higher in AD patient brain grey matter than in that of the controls (range +6 to 25%). The greatest relative differences were in the hippocampus (mean +25%, range -3 to 70%, p = 0.007), cerebellum (mean +11%, range -0.6 to 24%, p = 0.001), parietal (mean +44%, range -2 to 102%, p = 0.001) and temporal lobes (mean +9%, range -1 to 16%, p < 0.001). DTI MD and L1 results showed similar relative differences. We found strong positive associations between regional NODDI FISO and DTI RD across all ROIs in the AD patients, particularly for the parietal lobe (r^2^ = 0.81, p < 0.001) and hippocampus (r^2^ = 0.98, p < 7.4 × 10^−9^. The effect sizes for DTI RD were lower than for NODDI FISO across all ROIs.

DTI FA was lower in grey matter of brains of AD patients compared to controls only in some ROIs (range -12 to +2%). The greatest relative differences were observed in the hippocampus (mean -12%, range -30 to 5%, p = 0.006) and thalamus (mean -6%, range -19 to +2%, p < 0.023) and caudate (mean -9%, range -21 to +2%, p = 0.006). DTI RD showed a nominal trend towards negative correlations with SV2A, particularly in the parietal lobe in AD patients (r^2^ = 0.22, p = 0.15) (Figure 4B).

### Longitudinal changes in AD brains

We re-scanned the AD group after 12-18 months. We found small global decreases in brain volume at follow up, with greater relative volume changes in the temporal lobe (−4%, p=0.009), hippocampus (−5%, p=0.01), parahippocampal gyrus (−3%, p=0.027), cingulate cortex (−3%, p=0.023) and cerebellum (−2%, p=0.020). These were accompanied by small increases in FISO, significant in the medial temporal lobe (+5%, p = 0.046). We did not find any change in NDI and, although there were small reductions in mean ODI in the frontal lobe (−1%, p = 0.029) and in other ROIs, they were not statistically significant. We also found small and non-significant reductions in [^11^C]UCB-J DVR_CS-1_ after 12-18 months.

### Imaging associations with cognition

While our study was not well powered to test for relationships between the imaging and cognitive performance measures, we explored relationships amongst PET SV2A density and MRI microstructural imaging measures in the AD patients and controls. Lower regional SV2A was associated with impaired language (picture naming and semantic fluency) in the AD patients (hippocampus, r^2^ = 0.68, p=0.0001, parietal lobe, r^2^ = 0.39, p=0.03), but not in controls. Higher FISO in the parietal lobe was associated with impaired language in AD patients (r^2^ = 0.56, p=0.0084), but not in controls (r^2^ = 0.081, p=0.3) (Figure 5).

**Figure 5.**
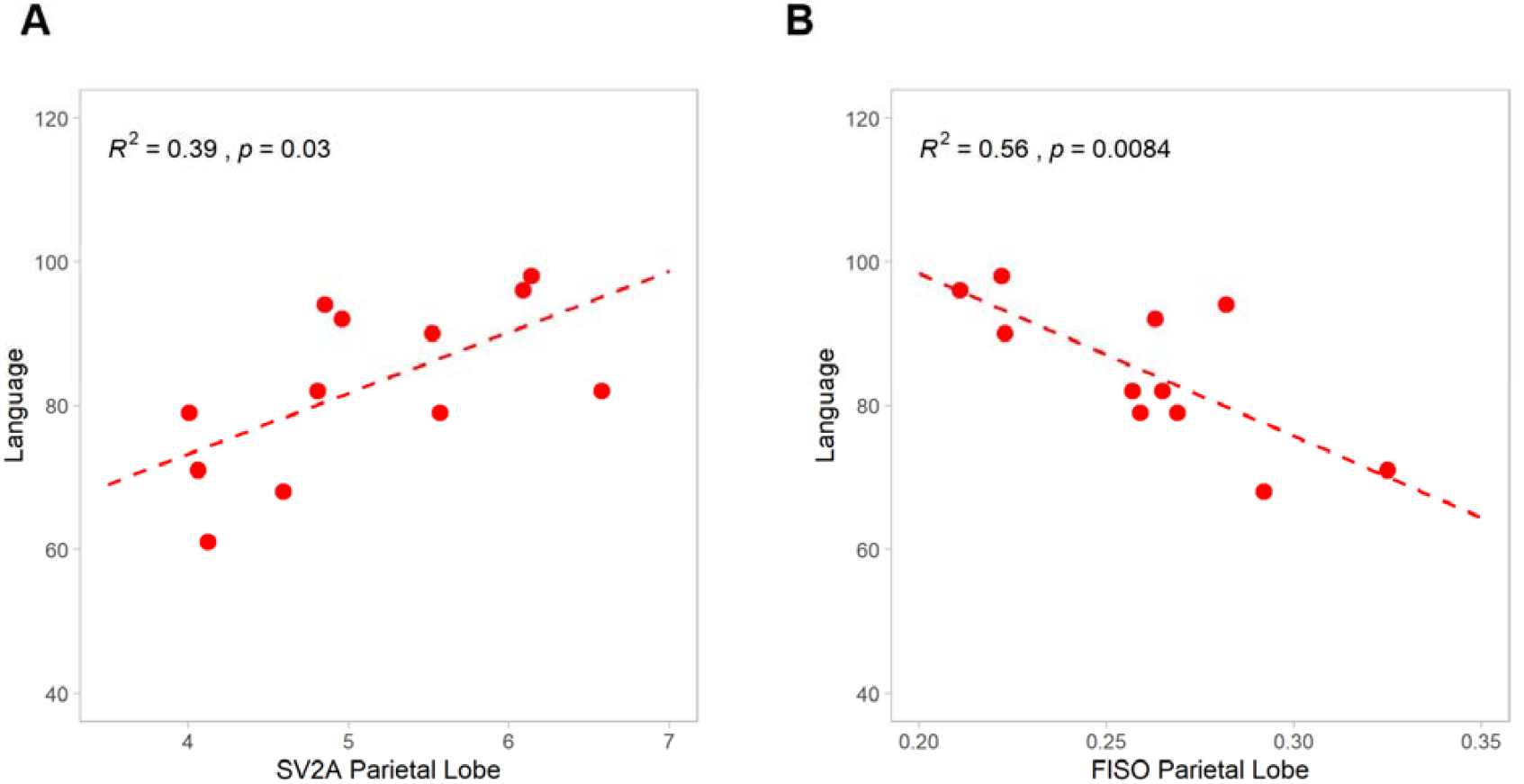
Loss of SV2A (A) and higher FISO (B) both were associated with impaired language (picture naming and semantic fluency) in the parietal lobe of AD patients

## DISCUSSION

We combined diffusion MRI and synaptic PET imaging to map changes in grey matter microstructure to characterise relationships between diffusion MRI measures and synaptic loss in early AD. Consistent with other recent reports, our study showed large decreases in SV2A density as well as brain atrophy in people with early AD. We also discovered striking increases in extracellular free water (FISO) in grey matter and consistent negative correlations between grey matter FISO and co-localised measures of both SV2A density and regional brain volume. FISO had a greater effect size for detecting differences between AD and controls in most ROIs than did the PET SV2A measures, regional volumes or the other NODDI and DTI indices; NDI and ODI showed only small relative changes in the AD brains relative to the healthy controls. We also found strong associations of FISO with DTI RD in the grey matter. Over 12-18 months of follow up of the AD patients, we found significant atrophy and FISO increases in the temporal lobe, although little change was measured in NDI, ODI or SV2A density. Exploration of relationships of the MRI and PET SV2A measures with cognitive performance suggested associations of higher parietal lobe FISO with lower language performance and greater SV2A with higher language performance.

SV2A is abundant in the brain, where it plays a role in presynaptic neurotransmitter release through Ca^2+^-dependent exocytosis [26] [27]. [^11^C]UCB-J was developed as a PET tracer to image the distribution and abundance of SV2A as an index of synaptic density [18,28,29]. Previous studies have shown substantial reductions of SV2A binding in medial temporal and neocortical brain regions in early AD [30–32]. We observed differences of similar sizes and distributions, even after correction of the [^11^C]UCB-J DVR for relative local tissue atrophy. [30,31]

Our most novel finding was of large increases in regional cortical FISO with AD that were inversely correlated with SV2A density. We interpret the increased FISO as an index of the glial activation[33,34]; glial water and the extracellular matrix changes together can modulate water diffusivity in the extracellular space. [35] Microglial and astrocyte activation are prominent in early AD. [7,36,37] Phagocytic microglial activation and pro-inflammatory astrocyte activation may be responsible for the pathological synaptic loss in AD. [38,39] An additional contribution may arise from changes in cortical mini-column organisation, particularly in later stages of AD. [40]

While there also may be a contribution from pathological loss of neuronal organisation, we believe that the small increases in neurite density and relative reductions in ODI, together with large FISO increases, are consistent with a model in which early AD neuropathology is dominated by synaptic loss, associated glial activation and a consequent reduction in the complexity of neuritic arborisation. While we observed small relative increases in neurite density (NDI), the latter could be a consequence of changes in neuronal organisation with distal neurite atrophy and relatively preserved proximal segments [41,42] or an adaptive increase in dendritic growth. [43]

The vast majority of diffusion imaging studies in AD have focused on white matter, [44,45]although changes in grey matter mean diffusivity have recently begun to be explored [33] Contributions of an explicitly-modelled free water compartment in AD have begun to be elucidated in the white matter, but the role of free water, and more sophisticated diffusion models more generally, have not previously been developed explicitly for grey matter. To date, NODDI has been applied in only a small number of human studies along the AD continuum, e.g., with healthy aging in the UK Biobank, [46] in mild cognitive impairment, [47,48] young onset AD [44,49,50], and sporadic late-onset AD. [45,51] Of these, only two have assessed grey matter changes in AD using NODDI and neither of these reported assessments of free water changes. One of these reports included data from people with early-onset, clinically defined AD that showed neurite density was lower in all ROIs compared to a healthy control group. Decreased ODI was found in inferior and middle temporal gyri, fusiform gyrus and precuneus, and cortical NDI was associated with cognitive impairment. [50] Another study explored correlations between grey matter changes in neurite density and tau PET in sporadic AD. [51] Decreased neurite density was associated regionally with tau deposition which is itself associated with neuroinflammation. Another, more recent study suggested FISO could provide an surrogate measure of tau deposition in AD. [52] Increased grey matter FISO was reported with a mouse model of tauopathy and in a recent human study of tau aggregate expression *in vivo*.[53] Although we found that the conventional DTI measures, which do not incorporate a microstructure compartmental model, also are sensitive to differences with disease (as reported by [40]), they have smaller effect sizes in the temporal lobe and hippocampus particularly, and have less specific pathological interpretations.

Despite limitations in size and scope, our study provides a foundation for characterising grey matter microstructure associated with synaptic loss and is the first to our knowledge to attempt this directly *in vivo*. While our study population was small, the combination of phenotyping depth with multi-modal imaging measures related to synaptic microstructure make it unique to our knowledge. The NODDI compartmental modelling appears informative but is interpreted using a simple model of grey matter structure and organisation that has not been validated quantitatively. This will demand new work well out of the scope of our investigations. Our study was not powered to test cognitive endpoints, so our tests for the clinical meaningfulness of the findings must be considered only as exploratory. Nonetheless, their general consistency with prior studies increases confidence in their potential generalisability. [30,33,52] Finally, although our AD patient and control groups were matched for sex, there was a 10 year mean age difference and both synaptic density and total GM volume decrease with aging. [54,55] However, age-related changes alone are relatively small [56]. In a larger cohort of 80 cognitively normal subjects (21-83 years old), the total brain GM volume was 2.9% lower per decade. Greatest age-related differences in GM SV2A density were reported in the caudate (6.4%/decade) [56,57]. Similar relative regional differences in GM SV2A density were found in the full MINDMAPS cohort of healthy controls (20-80 years old, n=24, unpublished), although age-related changes in SV2A binding measured were even lower (e.g., 3.5%/decade in the caudate). Group differences in SV2A binding and volume between AD and healthy controls that we observed for the current study were 3 to 7-fold greater than these values, so we do not believe that the age difference between our control and AD groups has a substantial impact on interpretation of our results.

## CONCLUSIONS

We have assessed the relationship between cortical microstructure and synapse loss *in vivo* in early AD for the first time. Our results suggest that NODDI MRI, and changes in free water in particular, could be more sensitive to differences in cortical pathology related to synaptic loss in early AD than the more molecularly-specific SV2A PET. Our observations suggest that NODDI MRI could be a useful adjunct to MRI measures of regional cortical volume change for stratification and monitoring of disease progression in people with AD.

## Data Availability

Non-identifible data produced in the present study are available upon reasonable request to the authors

## List of abbreviations

[^11^C]: 11-Carbon
[^18^F]: 18-Fluorine
1TC: One-Tissue Compartment
2TC: Two-Tissue Compartment
ACE III: Addenbrooke’s Cognitive Examination III
AD: Alzheimer’s Disease
AD: Axial Diffusivity
Aβ: Amyloid-β
CN: Cognitively Normal
CS: Centrum semiovale
DTI: Diffusion tensor imaging
DVR: Distribution volume ratio
ER: Endoplasmic Reticulum
ETC: Electron Transport Chain
FA: Fractional anisotropy
FISO: Fraction of isotropic
GM: Grey matter
MBq: Megabecquerel
MCI: Mild cognitive impairment
MD: Mean diffusivity
MMSE: Mini mental state examination
MNI: Montreal Neurological Institute
MRI: Magnetic Resonance Imaging
ms: Millisecond
mSv: Millisievert
NART: National Adult Reading Test
NDI: Neurite Density Index
NIA-AA: National Institute on Aging and Alzheimer’s Association
NODDI: Neurite Orientation Density and Dispersion Imaging
ODI: Orientation dispersion index
PET: Positron Emission Tomography
PVC: Partial Volume Correction
RD: Radial Diffusivity
ROI: Region of Interest
SPM12: Statistical Parametric Mapping version 12
SUVr: Standardised Uptake Value ratio
SV2A: Synaptic Vesicle Protein 2A
UCB-J: ((R)-1-((3-((11)C-methyl-(11)C)pyridin-4-yl)methyl)-4-(3,4,5-trifluorophenyl) pyrrolidin-2-one)
WM: White matter

## DECLARATIONS

### Ethics approval and consent to participate

All participants provided written informed consent. Ethical approval for this study was provided by the NHS London - Brighton & Sussex Research Ethics Committee (REC 18/LO/0179) for AD patients and East of England Cambridge Central & South Research Ethics Committee for controls. Radiation safety was approved by the Administration of Radioactive Substances Advisory Committee (ARSAC R92), London. Local site approval was provided by Imperial College London Joint Research Office.

### Authors’ contributions

Conceptualization: AVV, ALH, MH, JP, JBR, HT, DJB, LM, RAC, LC, AJS, RH, RNG, EAR and PMM

Methodology: CB, AM, GR, RNG, EAR and PMM

Investigation: AVV

Study regulation, recruitment: AVV,YL,EK

Formal Analysis: AVV

Visualization: AVV, CB

Funding acquisition: EAR,PMM

Supervision: PMM

Writing original draft: AVV, PMM

Writing review & editing: All authors

## Acknowledgements

We would like to thank all participants and their carers, patient and public involvement (PPI) groups and the Alzheimer’s Society Research Network Monitors who helped with this study. We would like to acknowledge the excellent and invaluable technical and administrative support provided by the Invicro-London radiochemistry, medical physics, blood laboratory, medical, nursing, PET technologist and both PET and MR radiography teams, to enable the collection of our data. A.V.V. is supported by the Alzheimer’s Society, grant number 440 (AS-CTF-18-006) and has received support from the NIHR Biomedical Research Centre at Imperial College London. P.M.M. acknowledges generous personal and research support from the Edmond J Safra Foundation and Lily Safra, an NIHR Senior Investigator Award, the UK Dementia Research Institute and the NIHR Biomedical Research Centre at Imperial College London. JBR is supported by the NIHR Cambridge Biomedical Research Centre (BRC-1215-20014) and Wellcome Trust (220258). This work is supported by the UK Dementia Research Institute which receives its funding from UK DRI Ltd, funded by the UK Medical Research Council, Alzheimer’s Society and Alzheimer’s Research UK. The views expressed are those of the authors and not necessarily those of the NIHR or the Department of Health and Social Care.

## REFERENCES

1. Prince M, Comas-Herrera A, Knapp M, Guerchet M, Karagiannidou M. World Alzheimer Report 2016 Improving healthcare for people living with dementia. Coverage, Quality and costs now and in the future. Alzheimer’s Dis. Int. [Internet]. 2016;1–140. Available from: https://www.alz.co.uk/research/world-report-2016

2. Livingston G, Huntley J, Sommerlad A, Ames D, Ballard C, Banerjee S, et al. Dementia prevention, intervention, and care: 2020 report of the Lancet Commission. Lancet. 2020;396:413–46.

3. Hardy J, Allsop D. Amyloid deposition as the central event in the aetiology of Alzheimer’s disease. Trends Pharmacol. Sci. 1991;12:383–8.

4. Hardy JA, Higgins GA. Alzheimer’s disease: The amyloid cascade hypothesis. Science (80-.). 1992;256:184–5.

5. Selkoe DJ. The molecular pathology of Alzheimer’s disease. Neuron. 1991;6:487–98.

6. Heneka MT, Golenbock DT, Latz E. Innate immunity in Alzheimer ‘ s disease. Nat. Immunol. [Internet]. Nature Publishing Group. 2015;16:229–36. Available from: http://dx.doi.org/10.1038/ni.3102

7. Leng F, Edison P. Neuroinflammation and microglial activation in Alzheimer disease: where do we go from here? Nat. Rev. Neurol. [Internet]. Springer US. 2021;17:157–72. Available from: http://dx.doi.org/10.1038/s41582-020-00435-y

8. DeKosky ST, Scheff SW. Synapse loss in frontal cortex biopsies in Alzheimer’s disease: Correlation with cognitive severity. Ann. Neurol. 1990;27:457–64.

9. Terry RD, Masliah E, Salmon DP, Butters N, Deteresa R, Hill R, et al. Physical Basis of Cognitive Alterations in Alzheimer ‘ s Disease[: Synapse h s s Is the Major Correlate of Cognitive Impairment. 1991;572–80.

10. Masliah E, Terry R. The Role of Synaptic Proteins in the Pathogenesis of Disorders of the Central Nervous System. Brain Pathol. 1993;3:77–85.

11. Coleman MP, Perry VH. Axon pathology in neurological disease: A neglected therapeutic target. Trends Neurosci. 2002. p. 532–7.

12. Serrano-Pozo A, Frosch MP, Masliah E, Hyman BT. Neuropathological alterations in Alzheimer disease. Cold Spring Harb. Perspect. Med. 2011;1:1–24.

13. Young PNE, Estarellas M, Coomans E, Srikrishna M, Beaumont H, Maass A, et al. Imaging biomarkers in neurodegeneration: Current and future practices. Alzheimer’s Res. Ther. Alzheimer’s Research & Therapy; 2020;12:1–17.

14. Scheltens P, Leys D, Barkhof F, Huglo D, Weinstein HC, Vermersch P, et al. Atrophy of medial temporal lobes on MRI in “ probable “ Alzheimer ‘ s disease and normal ageing[: diagnostic value and neuropsychological correlates. 1992;8:967–72.

15. Mak E, Holland N, Jones PS, Savulich G, Low A, Malpetti M, et al. In vivo coupling of dendritic complexity with presynaptic density in primary tauopathies. Neurobiol. Aging [Internet]. Elsevier Inc.; 2021;101:187–98. Available from: https://doi.org/10.1016/j.neurobiolaging.2021.01.016

16. Zhang Y, Fox GB. PET imaging for receptor occupancy: Meditations on calculation and simplification. J. Biomed. Res. 2012;26:69–76.

17. Südhof TC. Neuroligins and neurexins link synaptic function to cognitive disease. Nature [Internet]. 2008;455:903–11. Available from: http://www.pubmedcentral.nih.gov/articlerender.fcgi?artid=2673233&tool=pmcentrez&rendertype=abstract

18. Finnema SJ, Nabulsi NB, Eid T, Detyniecki K, Lin S, Chen M-K, et al. Imaging synaptic density in the living human brain. Sci. Transl. Med. [Internet]. 2016 [cited 2017 Jul 19];8. Available from: http://stm.sciencemag.org/content/8/348/348ra96.full

19. Rabiner EA. Imaging Synaptic Density: A Different Look at Neurologic Diseases. J. Nucl. Med. 2018;59:380–1.

20. Mansur A, Rabiner EA, Comley RA, Lewis Y, Middleton LT, Huiban M, et al. Characterization of 3 PET Tracers for Quantification of Mitochondrial and Synaptic Function in Healthy Human Brain: 18 F-BCPP-EF, 11 C-SA-4503, and 11 C-UCB-J. J. Nucl. Med. [Internet]. 2020;61:96–103. Available from: http://jnm.snmjournals.org/lookup/doi/10.2967/jnumed.119.228080

21. Wilson H, Pagano G, Natale ER De, Mansur A. Mitochondrial Complex 1, Sigma 1, and Synaptic Vesicle 2A in Early Drug-Naive Parkinson ‘ s Disease. 2020;1–12.

22. Whittington A, Gunn RN. Amyloid load: A more sensitive biomarker for amyloid imaging. J. Nucl. Med. 2019;60:536–40.

23. Tziortzi AC, Searle GE, Tzimopoulou S, Salinas C, Beaver JD, Jenkinson M, et al. Imaging dopamine receptors in humans with [11C]-(+)-PHNO: Dissection of D3 signal and anatomy. Neuroimage. 2011;54:264–77.

24. Müller-Gärtner HW, Links JM, Prince JL, Bryan RN, McVeigh E, Leal JP, et al. Measurement of radiotracer concentration in brain gray matter using positron emission tomography: MRI-based correction for partial volume effects. J. Cereb. Blood Flow Metab. 1992;12:571–83.

25. Smith SM, Jenkinson M, Woolrich MW, Beckmann CF, Behrens TEJ, Johansen-Berg H, et al. Advances in functional and structural MR image analysis and implementation as FSL. Neuroimage. 2004;23:208–19.

26. Vogl C, Tanifuji S, Danis B, Daniels V, Foerch P, Wolff C, et al. Synaptic vesicle glycoprotein 2A modulates vesicular release and calcium channel function at peripheral sympathetic synapses. Eur. J. Neurosci. [Internet]. John Wiley & Sons, Ltd (10.1111); 2015 [cited 2019 Jun 26];41:398–409. Available from: http://doi.wiley.com/10.1111/ejn.12799

27. Bajjalieh SM, Frantz GD, Weimann JM, McConnell SK, Scheller RH. Differential expression of synaptic vesicle protein 2 (SV2) isoforms. J. Neurosci. [Internet]. Society for Neuroscience. 1994 [cited 2019 Jun 26];14:5223–35. Available from: http://www.ncbi.nlm.nih.gov/pubmed/8083732

28. Nabulsi NB, Mercier J, Holden D, Carré S, Najafzadeh S, Vandergeten M-C, et al. Synthesis and Preclinical Evaluation of 11C-UCB-J as a PET Tracer for Imaging the Synaptic Vesicle Glycoprotein 2A in the Brain. J. Nucl. Med. [Internet]. Society of Nuclear Medicine. 2016 [cited 2017 Jul 19];57:777–84. Available from: http://www.ncbi.nlm.nih.gov/pubmed/26848175

29. Finnema SJ, Nabulsi NB, Mercier J, Lin SF, Chen MK, Matuskey D, et al. Kinetic evaluation and test–retest reproducibility of [11C]UCB-J, a novel radioligand for positron emission tomography imaging of synaptic vesicle glycoprotein 2A in humans. J. Cereb. Blood Flow Metab. [Internet]. SAGE PublicationsSage UK: London, England; 2018 [cited 2019 Jun 26];38:2041–52. Available from: http://journals.sagepub.com/doi/10.1177/0271678X17724947

30. Mecca AP, Chen M, O’Dell RS, Naganawa M, Toyonaga T, Godek TA, et al. In vivo measurement of widespread synaptic loss in Alzheimer’s disease with SV2A PET. Alzheimer’s Dement. [Internet]. 2020;16:974–82. Available from: https://onlinelibrary.wiley.com/doi/abs/10.1002/alz.12097

31. Chen M-K, Mecca AP, Naganawa M, Finnema SJ, Toyonaga T, Lin S, et al. Assessing Synaptic Density in Alzheimer Disease With Synaptic Vesicle Glycoprotein 2A Positron Emission Tomographic Imaging. JAMA Neurol. [Internet]. 2018 [cited 2018 Aug 28]; Available from: http://archneur.jamanetwork.com/article.aspx?doi=10.1001/jamaneurol.2018.1836

32. O’Dell RS, Mecca AP, Chen MK, Naganawa M, Toyonaga T, Lu Y, et al. Association of Aβ deposition and regional synaptic density in early Alzheimer’s disease: a PET imaging study with [11C]UCB-J. Alzheimer’s Res. Ther. Alzheimer’s Research & Therapy; 2021;13:1–12.

33. Montal V, Vilaplana E, Alcolea D, Pegueroles J, Pasternak O, González-Ortiz S, et al. Cortical microstructural changes along the Alzheimer’s disease continuum. Alzheimer’s Dement. 2018;14:340–51.

34. Weston PSJ, Simpson IJA, Ryan NS, Ourselin S, Fox NC. Diffusion imaging changes in grey matter in Alzheimer’s disease: A potential marker of early neurodegeneration. Alzheimer’s Res. Ther. [Internet]. Alzheimer’s Research & Therapy; 2015;7:1–8. Available from: http://dx.doi.org/10.1186/s13195-015-0132-3

35. Syková E, Nicholson C. Diffusion in brain extracellular space. Physiol. Rev. 2008;88:1277–340.

36. Calsolaro V, Matthews PM, Donat CK, Livingston NR, Femminella GD, Guedes SS, et al. Astrocyte reactivity with late-onset cognitive impairment assessed in vivo using 11C-BU99008 PET and its relationship with amyloid load. Mol. Psychiatry [Internet]. 2021; Available from: http://www.nature.com/articles/s41380-021-01193-z

37. Edison P, Archer HA, Gerhard A, Hinz R, Pavese N, Turkheimer FE, et al. Microglia, amyloid, and cognition in Alzheimer’s disease: An [11C](R)PK11195-PET and [11C]PIB-PET study. Neurobiol. Dis. [Internet]. Elsevier Inc.; 2008;32:412–9. Available from: http://dx.doi.org/10.1016/j.nbd.2008.08.001

38. Keren-Shaul H, Spinrad A, Weiner A, Matcovitch-Natan O, Dvir-Szternfeld R, Ulland TK, et al. A Unique Microglia Type Associated with Restricting Development of Alzheimer’s Disease. Cell [Internet]. Elsevier. 2017;169:1276-1290.e17. Available from: http://dx.doi.org/10.1016/j.cell.2017.05.018

39. Bartels T, De Schepper S, Hong S. Microglia modulate neurodegeneration in Alzheimer’s and Parkinson’s diseases. Science (80-.). 2020;370:66–9.

40. Torso M, Bozzali M, Zamboni G, Jenkinson M, Chance SA. Detection of Alzheimer’s Disease using cortical diffusion tensor imaging. Hum. Brain Mapp. 2021;42:967–77.

41. Cotman CW, Matthews DA, Taylor D, Lynch G. Synaptic rearrangement in the dentate gyrus: histochemical evidence of adjustments after lesions in immature and adult rats. Proc. Natl. Acad. Sci. U. S. A. 1973;70:3473–7.

42. Scheff S. Reactive Synaptogenesis in Aging and Alzheimer’s Disease: Lessons Learned in the Cotman Laboratory. Neurochem. Res. 2003;28:1625–30.

43. Jackson JS, Witton J, Johnson JD, Ahmed Z, Ward M, Randall AD, et al. Altered Synapse Stability in the Early Stages of Tauopathy. Cell Rep. [Internet]. ElsevierCompany.; 2017;18:3063–8. Available from: http://dx.doi.org/10.1016/j.celrep.2017.03.013

44. Slattery CF, Zhang J, Paterson RW, Foulkes AJM, Carton A, Macpherson K, et al. ApoE influences regional white-matter axonal density loss in Alzheimer’s disease. Neurobiol. Aging [Internet]. Elsevier Inc. 2017;57:8–17. Available from: http://dx.doi.org/10.1016/j.neurobiolaging.2017.04.021

45. Raghavan S, Reid RI, Przybelski SA, Lesnick TG, Schwarz CG, Knopman DS, et al. Diffusion models reveal white matter microstructural changes with aging, pathology, and cognition. 2021;1–42.

46. Cox SR, Ritchie SJ, Tucker-Drob EM, Liewald DC, Hagenaars SP, Davies G, et al. Ageing and brain white matter structure in 3,513 UK Biobank participants. Nat. Commun. [Internet]. Nature Publishing Group. 2016;7:1–13. Available from: http://dx.doi.org/10.1038/ncomms13629

47. Wen Q, Mustafi SM, Li J, Risacher SL, Tallman E, Brown SA, et al. White matter alterations in early-stage Alzheimer’s disease: A tract-specific study. Alzheimer’s Dement. Diagnosis, Assess. Dis. Monit. 2019;11:576–87.

48. Fu X, Shrestha S, Sun M, Wu Q, Luo Y, Zhang X, et al. Microstructural White Matter Alterations in Mild Cognitive Impairment and Alzheimer’s Disease: Study Based on Neurite Orientation Dispersion and Density Imaging (NODDI). Clin. Neuroradiol. 2020;30:569–79.

49. Parker CA, Nabulsi N, Holden D, Lin S -f., Cass T, Labaree D, et al. Evaluation of 11C-BU99008, a PET Ligand for the Imidazoline2 Binding Sites in Rhesus Brain. J. Nucl. Med. [Internet]. 2014;55:838–44. Available from: http://jnm.snmjournals.org/cgi/doi/10.2967/jnumed.113.131854

50. Parker TD, Slattery CF, Zhang J, Nicholas JM, Paterson RW, Foulkes AJM, et al. Cortical microstructure in young onset Alzheimer’s disease using neurite orientation dispersion and density imaging. Hum. Brain Mapp. 2018;39:3005–17.

51. Sone D, Shigemoto Y, Ogawa M, Maikusa N, Okita K, Takano H, et al. Association between neurite metrics and tau/inflammatory pathology in Alzheimer’s disease. Alzheimer’s Dement. Diagnosis, Assess. Dis. Monit. 2020;12:1–10.

52. Anderson, J; Schifani, C; Nazeri, A; Volneskos A for the ADNI. In Vivo Cortical Microstructure: A Proxy for Tauopathy and Cognitive impairment in the Elderly with and without MCI/Dementia. medRxiv. 2021;8:1–13.

53. Colgan N, Siow B, O’Callaghan JM, Harrison IF, Wells JA, Holmes HE, et al. Application of neurite orientation dispersion and density imaging (NODDI) to a tau pathology model of Alzheimer’s disease. Neuroimage [Internet]. The Authors. 2016;125:739–44. Available from: http://dx.doi.org/10.1016/j.neuroimage.2015.10.043

54. Peter R. H. Synaptic density in human frontal cortex -Developmental changes and effects of aging. Brain Res. 1979;163:195–205.

55. Hutton C, Draganski B, Ashburner J, Weiskopf N. A comparison between voxel-based cortical thickness and voxel-based morphometry in normal aging. Neuroimage [Internet]. Elsevier Inc.; 2009;48:371–80. Available from: http://dx.doi.org/10.1016/j.neuroimage.2009.06.043

56. Toyonaga, T, Lu Y, Naganawa, M, Matuskey, D, Mecca, A, Nabulsi, NB, Chen, M,, Esterlis, I, Van Dyck, CH, Huang, Y, Carson R. Relationship of Age and Synaptic Density: A 11C[UCB[J PET Study in Healthy Controls with Partial Volume Correction. SNMMI Oral Presentation. 2019.

57. Toyonaga T, Lu Y, Naganawa M, Matuskey D, Mecca A, Pittman B, et al. Relationship of Age and Synaptic Density:A 11C-UCB-J PET Study in Healthy Controls with Partial Volume Correction. J. Nucl. Med. [Internet]. 2019;60:423 LP –423. Available from: http://jnm.snmjournals.org/content/60/supplement_1/423.abstract

